# Diagnostic and prognostic value of the gasdermins in gastric cancer

**DOI:** 10.1101/2023.10.18.23297225

**Authors:** Yeqiong Xu, Chuandan Wan, Ping Wang, yulan Gu

## Abstract

**Background:** Pyroptosis has been drawn attention owing to its contributory role in various cancers. Recently, the participator of pyroptosis, gasdermins (GSDMs) have been reported associated with of multiple types of cancers. However, the role of GSDMs expression in diagnosis and prognosis of gastric cancer (GC) has not been well elucidated. Moreover, the mechanisms underlying the carcinogenesis of GC are still obscure.

**Methods:** Herein we analyzed the transcriptional, prognostic information and the role of GSDMs in patients with GC from TIMER, UALCAN, Human Protein Atlas (HPA), GEPIA and Kaplan-Meier plotter databases. The cBioPortal online tool was used to analyze the GSDMs alterations, correlations, and networks. Furthermore, String, Cytoscape and TIMER were conducted to explore the functional enrichment and immune modulation. The statistical analysis was carried out in the R environment, and *P*-value < 0.05 was considered statistically significant.

**Results:** GSDMB, GSDMC, GSDMD, GSDME were with higher expression in GC than normal tissue in TIMER database. Moreover, survival analyses via two databases both demonstrated that high expression of GSDME was related to shorter overall survival (OS) in patients with GC. Additionally, functional enrichment revealed that GSDMs might be involved in endopeptidase activity, peptidase regulator activity, cysteine-type peptidase activity. Besides, GSDMs were correlated with infiltration levels of immune cells in GC, and GSDME was correlated with the infiltrating level of CD4+ T, CD8+ T, neutrophils, macrophages and dendritic cells.

**Conclusions:** The study systematically indicated the potential diagnostic and prognostic value of GSDMs in GC. Our results showed that GSDME might play a considerably oncogenic role in GC diagnosis and prognosis. However, our bioinformatics analyses should be further validated in more prospective studies.

## Introduction

GC, one of the most prevalently malignant tumors, influences both the physical and mental health of those at risk, particularly in East Asia. It has been reported that an estimated 1,089,103 new GC cases and 768,793 GC related deaths worldwide in 2020, making GC sixth in cancer-specific incidence and mortality ranks third [1]. Although multimodal treatment strategies, including chemotherapy, radiotherapy, immunotherapy, and non-invasive surgical resection have greatly advanced in recent decades, the outcomes of curing GC remain unsatisfactory, and the 5-year survival rate for patients with GC remains unfavorable due to local relapse or distant metastasis [2], causing a major burden on families and society. Therefore, new biomarkers as molecular pathology diagnosis and prognosis prediction to effectively improve prognosis and individualized treatment are urgent needed.

Pyroptosis is a proinflammatory process which features plasma membrane rupture due to gasdermin pore formation [3–6]. As critical inductions to execute pyroptosis [5], the gasdermins (GSDMs) are consisted of a family of pore-forming proteins, GSDMA, GSDMB, GSDMC, GSDMD, GSDME (DFNA5) and PJVK (DFNB59), which all display differential tissue expression [6]. Recently, GSDMs have been reported involved in a diversity of cellular activities, such as inflammation, cell proliferation, differentiation and cell death [7, 8], which indicated that GSDMs were related to various oncological pathologies, such as GC [9, 10], breast cancer [11], colorectal cancer [12], lung cancer [13]. Although GSDMs were related to patients with GC, its specific role and associated tumorigenesis mechanisms remain unclear. Therefore, an in-depth investigation of the association between GSDMs and GC will provide new directions and targets for the detection, treatment and prevention of GC.

The present study was carried out to investigate the detailed function and molecular mechanism of the GSDMs in GC biology. By utilizing TIMER, UALCAN and HPA databases, the transcriptional levels and protein expression of GSDMs were measured. The cBioPortal online tool was used to analyze the GSDMs alterations, correlations, and networks. As for the prognosis of patients with GC, the Kaplan-Meier plotter and GEPIA were used. Furthermore, Gene Ontology functions and biological pathways of GSDMs together with their related genes were predicted via STRING, Cytoscape Databases. Our study presents the potential biological function and prognostic value of GSDMs, which will broaden the horizon of diagnosis and treatment of GC.

## Materials and Methods

### TIMER Database Analysis

TIMER 2.0 (http://timer.comp-genomics.org/) is applied to analyze the expression levels of GSDMs in various normal or tumor cells and the infiltration levels of immune cells in 31 tumor types [14]. The scatterplots of GSDMs were generated to show the purity-corrected partial Spearman’s rho value and statistical significance. The positive purity value expected genes are highly expressed in the tumor cells, and the opposite is expected for genes highly expressed in the microenvironment. In the current study, we analyzed the correlation between the expression of GSDMs with the abundances of six tumor-infiltrating immune cell (B cells, CD8+ T cells, CD4+ T cells, neutrophils, dendritic cells and macrophages).

### UALCAN database

UALCAN database a publicly accessible online database, was applied to analyze the mRNA expression of GSDMs in GC based on the OMICS (including TCGA, MET500, and CPTAC databases) [15]. Furthermore, this database was also analyzed the mRNA expression of GSDMs in GC stage and grade subgroup.

### GEPIA Database Analysis

GEPIA (http://gepia.cancer-pku.cn/), a publicly accessible online database, was applied to analyze the prognosis value of GC patients according GSDMs expressions based on the GTEx and TCGA databases [16], with 9,736 and 8,587 tumor samples, respectively.

### Kaplan–Meier plotter

Kaplan–Meier plotter (http://kmplot.com/analysis/index.php?p=background), the publicly database, is used to analyze the association between different genes expressions and survival in diversity cancers. Herein, the database was conducted to explore the prognosis value in GC patients based on GSDMs expressions.

### HPA database Analysis

In addition to the UALCAN database providing mRNA expression analysis of GSDMs family members, HPA (http://www.proteinatlas.org/) as a platform containing representative immunohistochemistry-based protein expression data for near 20 highly common kinds of cancers [17]. Through this database, the levels of GSDMs protein expression were detected in GC and normal tissues.

### cBioPortal Database Analysis

The cBioPortal Database (https://www.cbioportal.org/results/oncoprint?data_priority=0&tab_index=tab_visualize&Action=Submit&session_id=607bb3d9e4b015b63e9e6853) was used to analyze mutations, putative copy number alterations (CNAs) from genomic identification of significant targets in cancer (GISTIC), mRNA expression Z scores (RNA-seq v.2 RSEM), and protein expression Z scores (reverse phase protein array [RPPA]). Co-expression and network were calculated according to the cBioPortal’s online instructions.

### STRING and Cytoscape

String is an online tool dedicated to the study of protein-protein interaction (PPI) relationships [18]. It provides data predictions including protein predictions, protein interactions, co-expression, sharing of protein domains, subcellular colocalization, signaling pathways, genetic interactions. Cytoscape is a publicly platform for network analysis and visualization. In the current study, we analyzed proteins interacting with members of the GSDMs family.

## Statistical analysis

The statistical analysis and graphical work were carried out in the R environment (Version 3.6.3). All statistical tests were bilateral, and *P*-value < 0.05 was considered statistically significant.

## Ethical Considerations

The present study was a retrospective study based on existing internet public data and did not require ethical approval. All the data were fully anonymized before accessed by our research team and the ethics committee waived the requirement for informed consent.

## Results

### Transcriptional Levels of GSDMs in distinct types of cancers

To investigate the critical role of GSDMs in carcinogenesis, the transcriptional levels of six members in gasdermin family among cancers were compared in various cancers with those in normal tissues by TIMER database (Figure 1). The results indicated that GSDMs were highly expressed in numerous tumors, including cholangiocarcinoma, uterine corpus endometrial carcinoma, bladder urothelial carcinoma and STAD. Among all types of cancers, GSDMB, GSDMC, GSDMD, GSDME were with higher expression in STAD than normal tissue.

**Figure 1.**
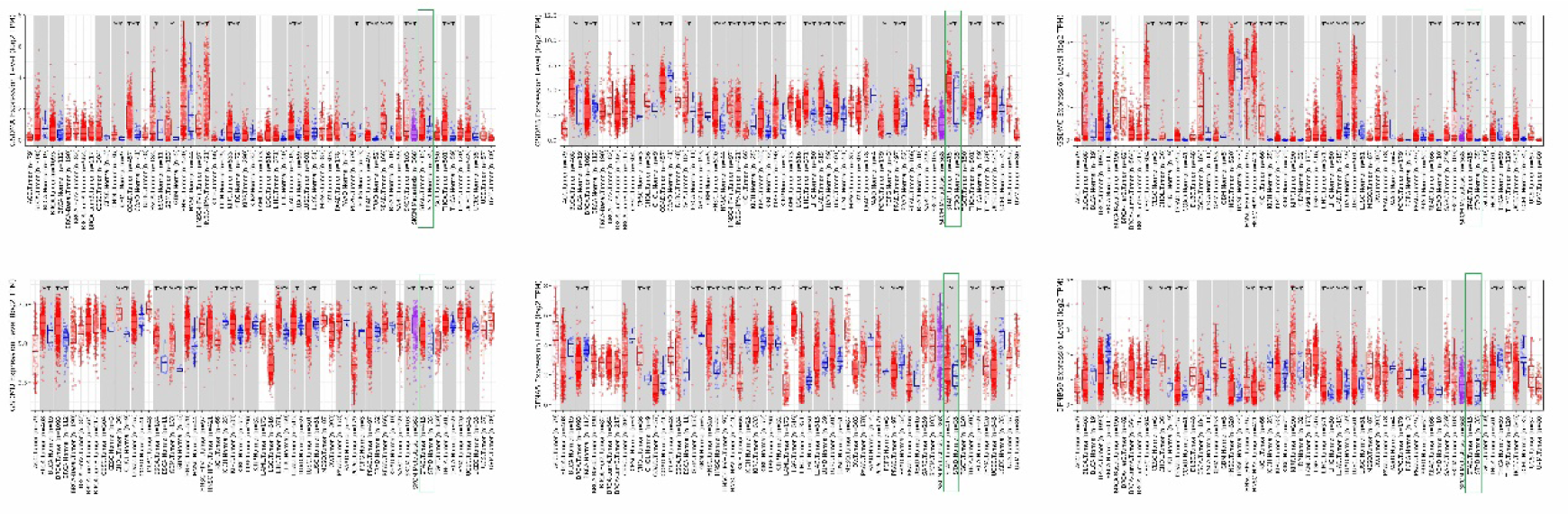
GSDMs expression levels in multiple types of tumor or normal cells via TIMER analysis (\**P*<0.05, \*\**P*<0.01, \*\*\**P*<0.001).

### Association between the expression of GSDMs and the clinicopathological parameters of patients with GC

The mRNA expression of GSDMs between GC and normal gastric tissues were compared by UALCAN database. The results showed that the levels of GSDMA, GSDMB, GSDMD and GSDME were higher in STAD than in normal tissues (Figure 2). Then the two key clinicopathological parameters of GC, tumor stage and tumor grade were further analyzed in the expression of GSDMs. The expressions of GSDMA, GSDMB, GSDMD and GSDME were significantly higher in tumor stage 2 and 3 than normal tissues. Meanwhile, the expressions of GSDMA and GSDMD were also higher in tumor stage 1, compared with that in normal tissues. Besides, all GSDMs (except GSDME) expressions had no difference between tumor stage 4 and normal tissues. (Figure 3A). In terms of tumor grade, the expression of GSDMD gradually increased from tumor grade 1 to 4 and higher than normal tissue. The expressions of GSDMB and GSDME were higher in tumor grade 2-3 compared to normal tissue, whereas GSDMA expression was significantly increased in tumor grade 2 compared to normal tissue. Besides, GSDMC and PJVK expressions differed in tumor grade 1 and 3 from normal tissue, respectively (shown in Figure 3B).

**Figure 2.**
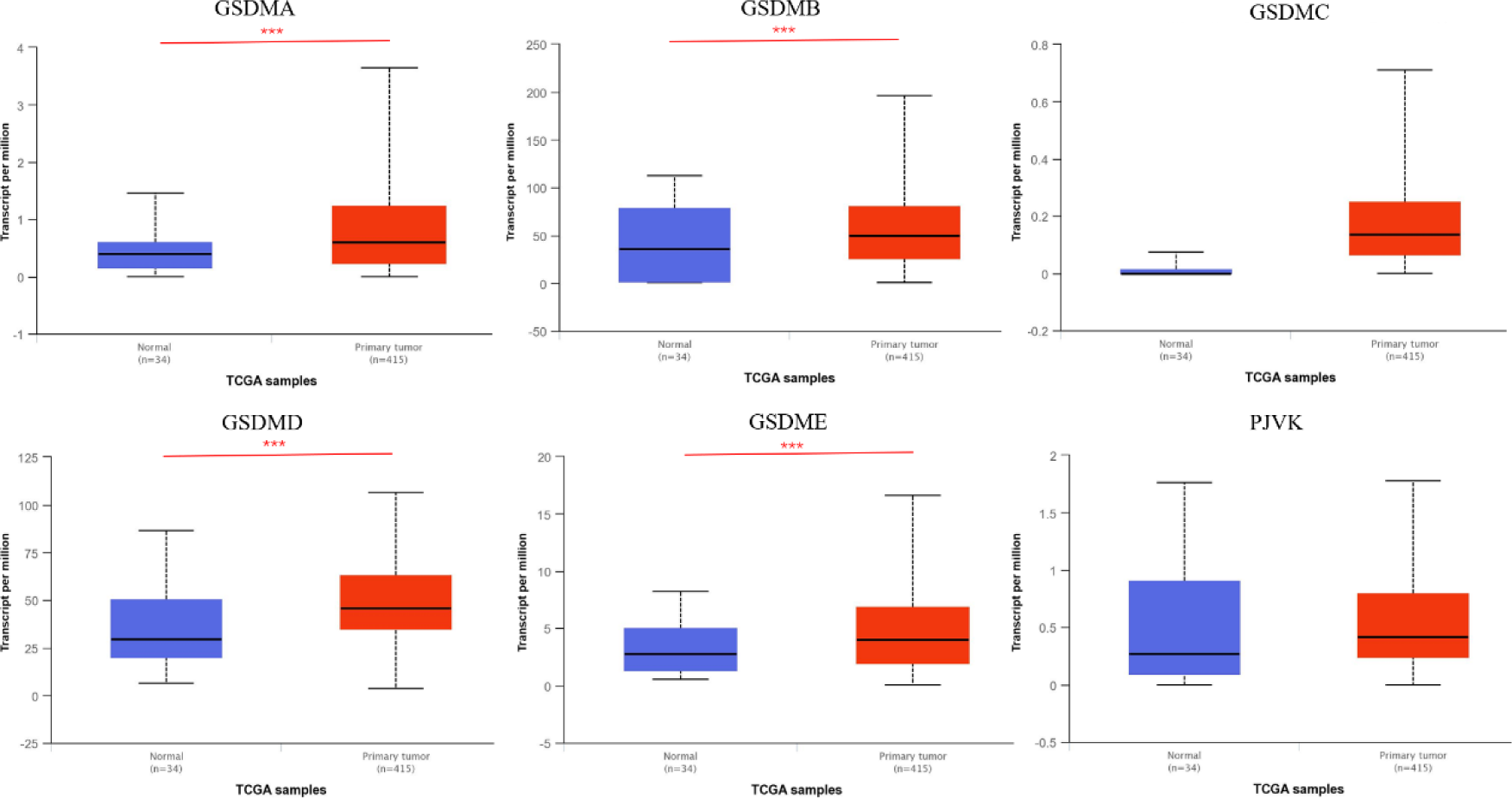
GSDMs expression levels in GC by UALCAN database (\**P*<0.05, \*\**P*<0.01, \*\*\**P*<0.001).

**Figure 3.**
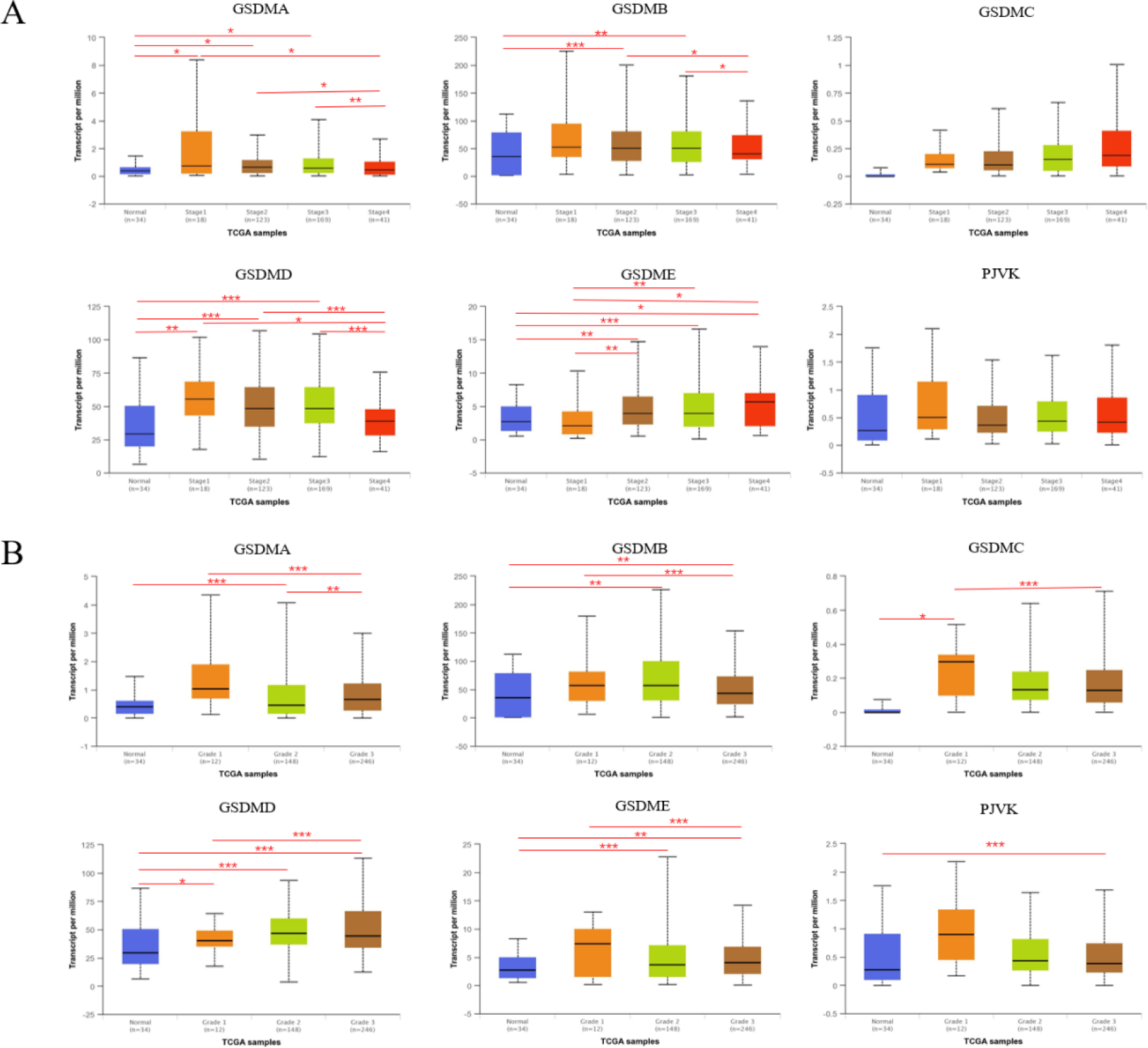
Correlation between GSDMs mRNA expression levels and clinicopathological parameters of patients with GC by UALCAN database (\**P*<0.05, \*\**P*<0.01, \*\*\**P*<0.001). (A) Correlation between GSDMs mRNA expression levels and individual cancer stages of GC. (B) Correlation between GSDMs mRNA expression levels and individual cancer grades of GC.

At the same time, we conducted immunohistochemistry (IHC) to compare the GSDMs expression in GC tissues and their counterparts by HPA (shown in Figure 4). The results indicated that GSDMA and GSDMD proteins were more highly expressed in the GC tissues than in the normal tissues, which was in line with the tendency of their mRNA expression. However, GSDMB, GSDMC and GSDME proteins showed no difference between GC tissues and their counterparts, and no IHC data of PJVK was retrieved from the THPA database.

**Figure 4.**
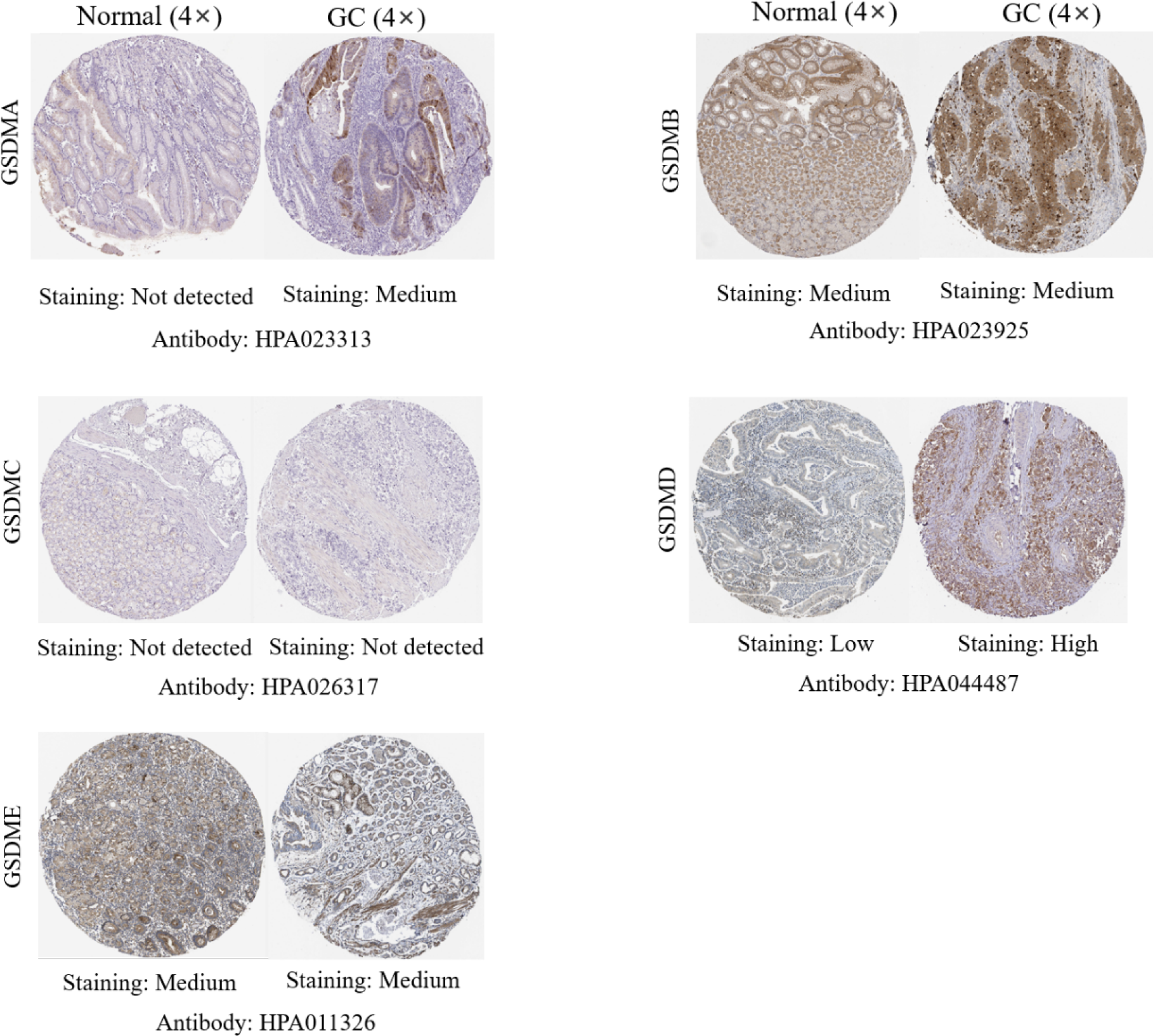
GSDMs expression in patients with GC using IHC (HPA).

### Relationship between the mRNA expression of GSDMs and prognosis of patients with GC

Furthermore, we investigated the key role of GSDMs in the survival of patients with GC. The results of Kaplan-Meier plotter and GEPIA appeared inconsistent. The former showed the high level of GSDMB, GSDMD, GSDME and PJVK was associated with worse OS in patients with GC (Figure 5A). However, no data of GSDMA was retrieved from the Kaplan-Meier plotter database. The latter revealed that only high expression of GSDME was related to shorter OS in patients with GC (Figure 5B).

**Figure 5.**
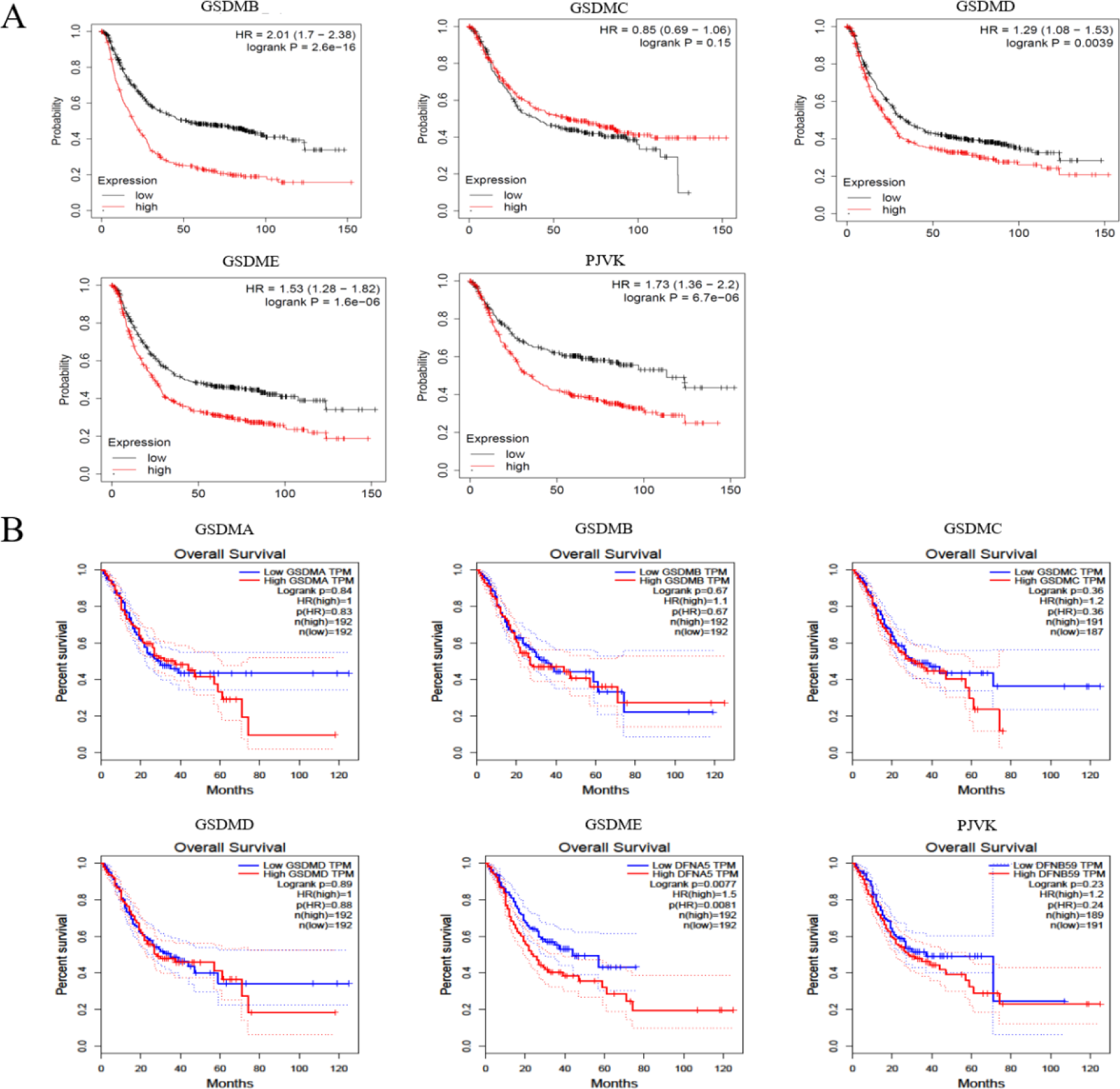
The prognostic value of GSDMs expression levels in patients with GC. (A) Correlation between GSDMs mRNA expression levels and OS of patients of GC by Kaplan-Meier plotter database. (B) Correlation between GSDMs mRNA expression levels and OS of patients of GC by GEPIA database.

### Predicted functions and pathways of the changes in GSDMs and their frequently altered neighbor genes in patients with GC

Then, we made a comprehensive identification of the GSDMs molecular features owing to their clinical characteristics. The cBioPortal online tool was used to analyze the GSDMs alterations, correlations, and networks. The alterations of GSDMs were detected in 187 samples (39%). The alteration frequency of GSDMA, GSDMB, GSDMC, GSDMD, GSDME and PJVK were 13%, 13%, 13%, 17%, 7% and 5% based on cBioPortal database (Figure 6A). In detail, amplification and mRNA high were the two main alteration types of genetic alteration of GSDMs. However, missense mutation, splice mutation, truncating mutation and deep deletion were rarely in GSDMs. Meanwhile, the homologous correlations of GSDMs were analyzed, showing that strong correlation in GSDMA and GSDMB (R=0.58), and weak correlation in PJVK and GSDMD (R=-0.21) (Figure 6B).

**Figure 6.**
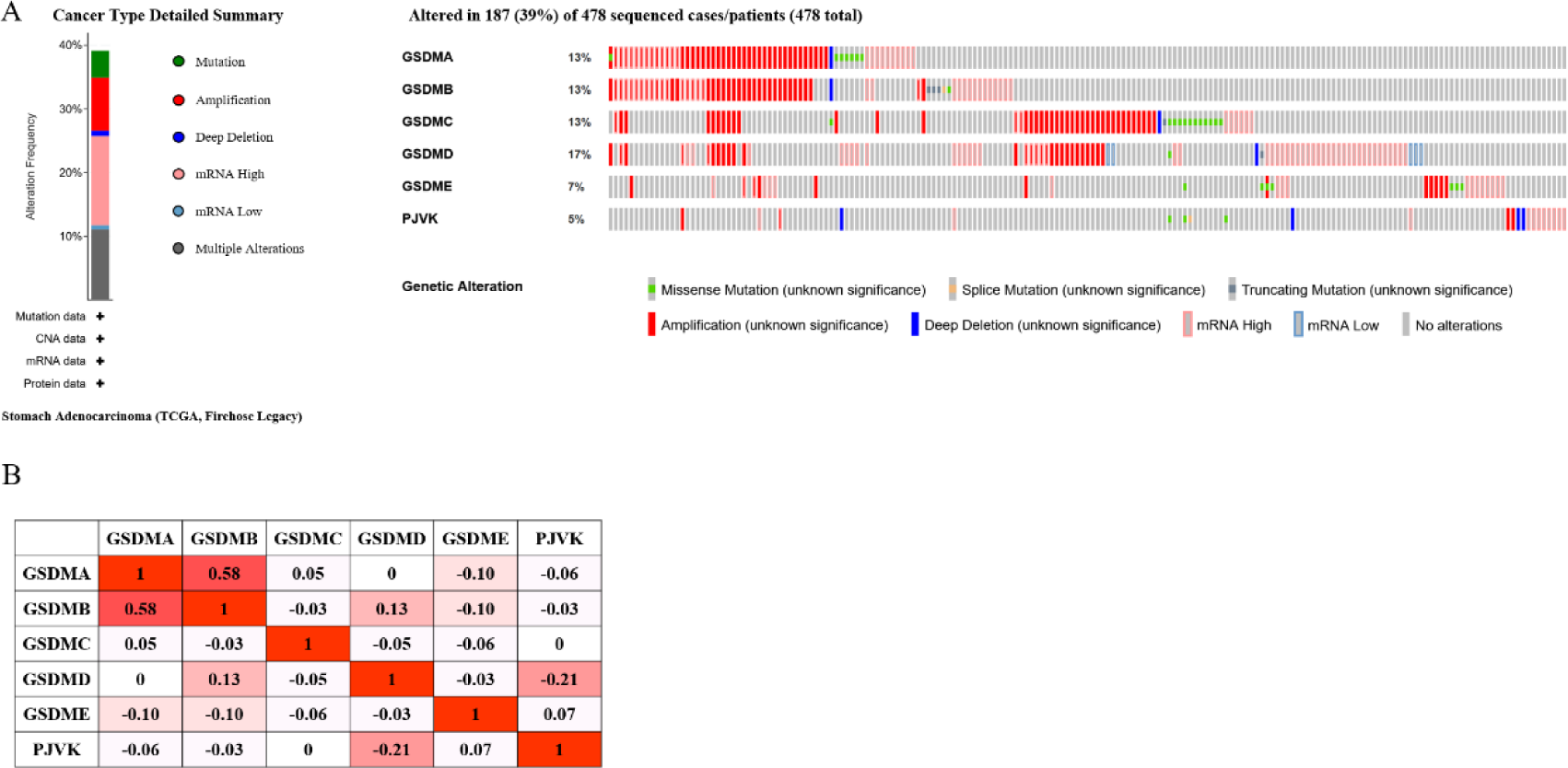
GSDMs gene expression and mutation analysis in GC using cBioPortal database. (A) GSDMs gene expression and mutation analysis in GC. (B) Correction between different GSDMs in GC.

Moreover, STRING and Cytoscape were used to screen out the genes which were co-expressed and associated with the GSDMs, showing that the physical interactions among the six genes were significant in this network (Figure 7A). Furthermore, Gene Ontology (GO) annotation and Kyoto Encyclopedia of Genes and Genomes (KEGG) pathway analyses were devoted to determining their biological function. The top-ranking biological processes regarding GSDMs were cytokine production, regulation of cysteine-type endopeptidase activity, regulation of proteolysis, regulation of inflammatory response and response to virus (Figure 7B). As for cellular components of GSDMs, the results focused on inflammasome complex, actin-based cell projection, cluster of actin-based cell projections, stereocilium bundle and stereocilium (Figure 7C). Moreover, the highly enriched molecular functions of GSDMs were endopeptidase activity, peptidase regulator activity, cysteine-type peptidase activity, cysteine-type endopeptidase activity involved in apoptotic process and peptidase activator activity (Figure 7D). The KEGG pathway analysis indicated that the top five were NOD-like receptor signaling pathway, Salmonella infection, Shigellosis, Pathogenic Escherichia coli infection and Influenza A (Figure 7E).

**Figure 7.**
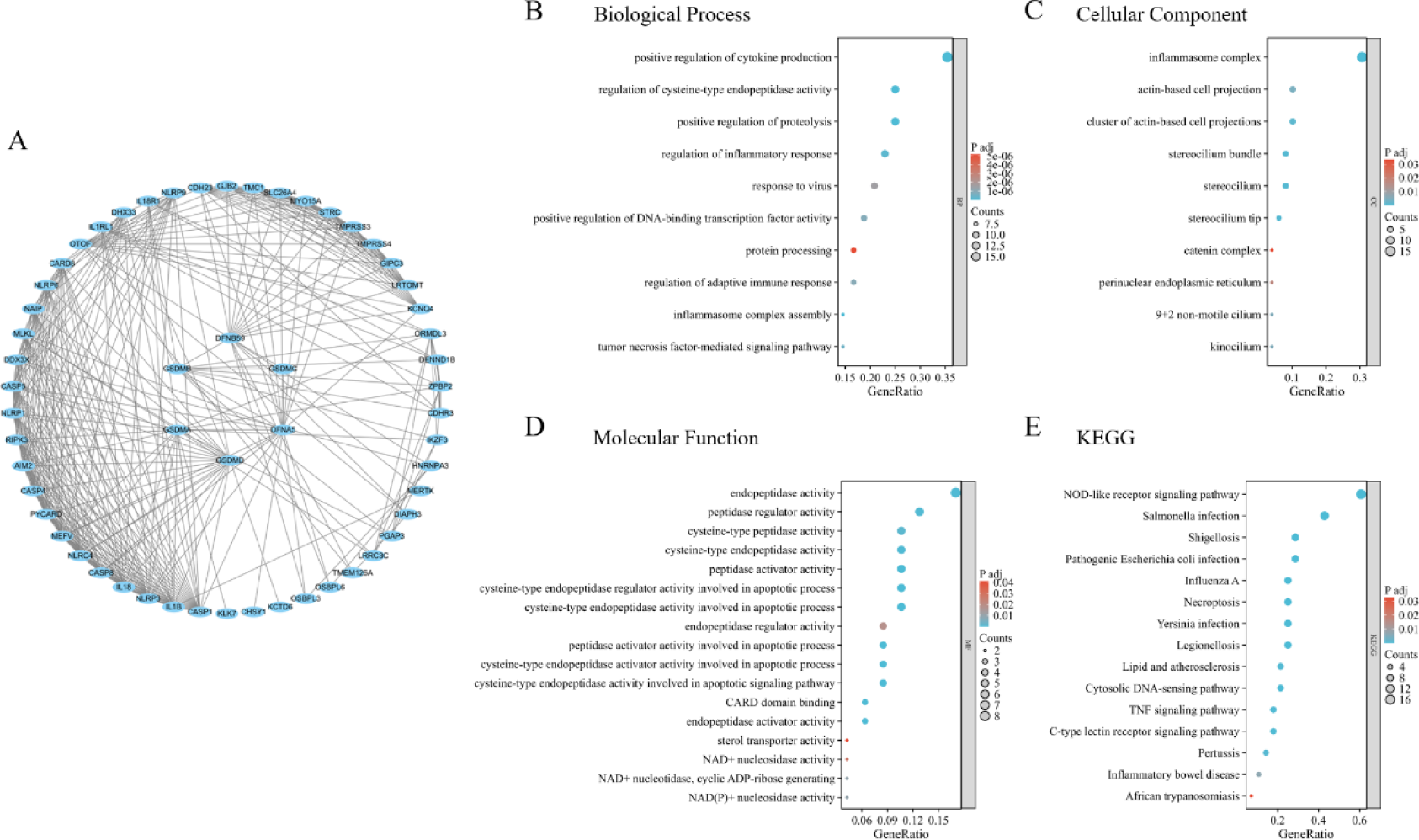
Function enrichment and pathways of GSDMs and co-expression genes in GC. (A) The network for GSDMs and the most frequently altered co-expressed genes in GC using STRING and visualization by Cytoscope. (B) Biological process analysis of GSDMs associated co-expressed genes. (C) Cellular component analysis of GSDMs associated co-expressed genes. (D) Molecular function analysis of GSDMs associated co-expressed genes. (E) KEGG pathway analysis of GSDMs associated co-expressed genes.

### Association of mRNA expression of GSDMs with immune infiltration level in TCGASTAD patients

Then, TIMER 2.0 was utilized to determine the correlation between the expression of GSDMs and infiltration levels of immune cells in STAD (Figure 8). From a point of view, the mRNA expression levels of GSDMC and GSDME were obviously related to tumor purity. More specifically, GSDMA was positively correlated with the infiltration of CD8+ T cells (*Rho*=0.178, *P*=5.17e-04), neutrophils (*Rho*=0.184, *P*=3.23e-04), dendritic cells (*Rho*=0.244, *P*=1.46e-06) and macrophages (*Rho*=0.166, *P*=1.15e-03). Moreover, GSDMB was positively correlated with the infiltration of B cells (*Rho*=0.123, *P*=1.64e-02), while was negatively correlated with the infiltration of macrophages (*Rho*=-0.360, *P*=4.51e-13). Similarly, a positive correlation was observed between GSDMC and the infiltration of CD8+ T cells (*Rho*=0.158, *P*=2.05e-03), dendritic cells (*Rho*=0.136, *P*=7.94e-03) and neutrophils (*Rho*=0.269, *P*=1.05e-07), while a negative correlation with CD4+ T cells infiltration (*Rho*=-0.305, *P*=1.34e-09). Likewise, a positive correlation was observed in the infiltration of B cells (*Rho*=-0.202, *P*=1.34e-09), CD4+ T cells (*Rho*=0.158, *P*=2.05e-03), dendritic cells (*Rho*=0.136, *P*=7.94e-03) with GSDMD expression level. Conversely, there was a negative correlation between GSDMD expression level and the infiltration of macrophages (*Rho*=-0.178, *P*=5.07e-04). CD4+ T cells (*Rho*=0.292, *P*=6.70e-09), CD8+ T cells (*Rho*=0.164, *P*=1.35e-03), neutrophils (*Rho*=0.166, *P*=1.14e-03), dendritic cells (Rho=0.224, *P*=1.07e-05) and macrophages (*Rho*=0.451, *P*=2.12e-20) all had a positive correlation with GSDME expression level. Finally, PJVK was positively correlated with the infiltration of macrophages (*Rho*=0.154, *P*=2.57e-03), while a negative correlation with dendritic cells infiltration (*Rho*=-0.132, *P*=1.00e-02).

**Figure 8.**
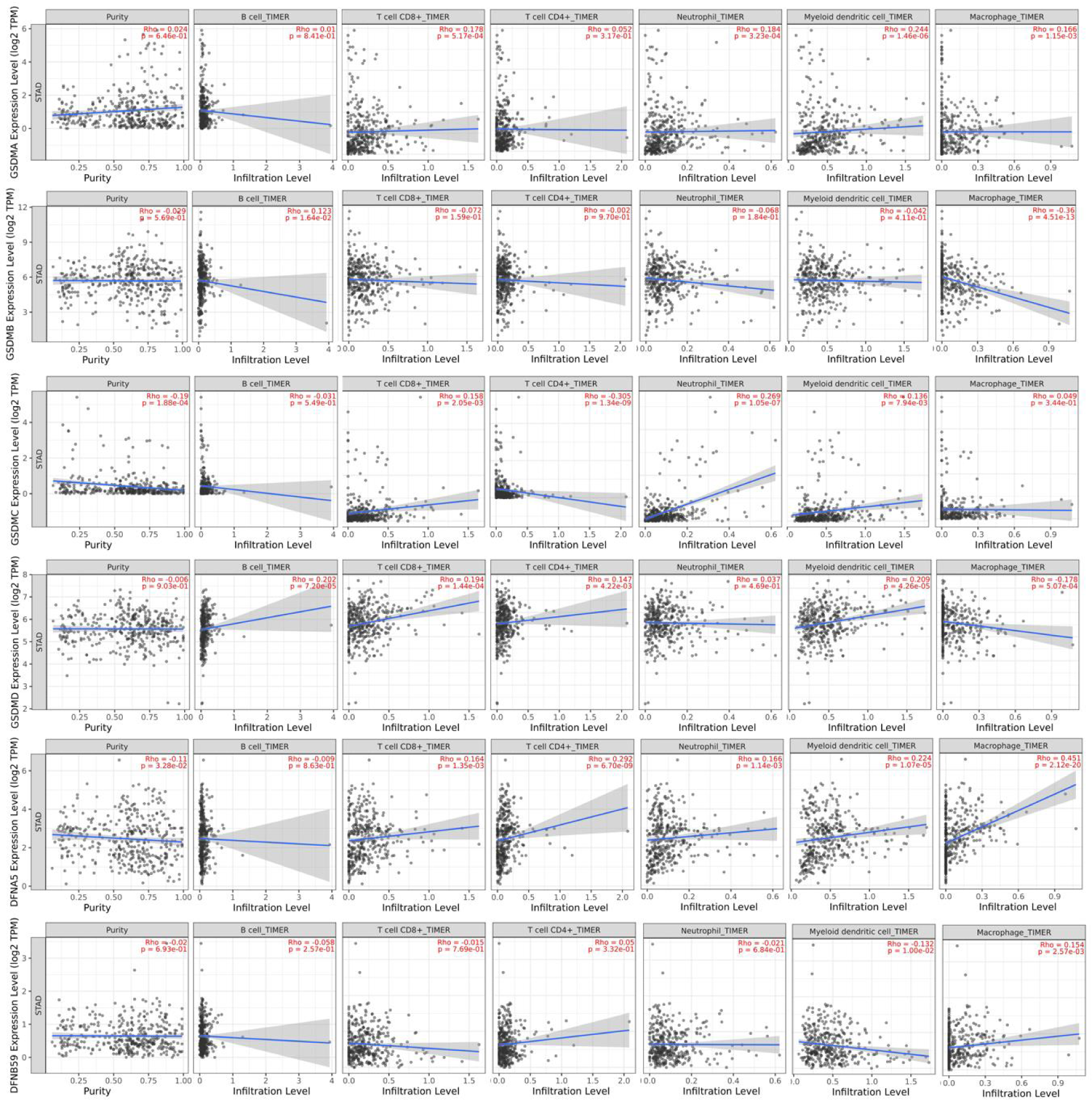
Correlation analysis between GSDMs expression and immune cell infiltration levels in GC by TIMER database.

## Discussion

GC remains one of the most important cancers worldwide with over 1,000,000 new cases and 783,000 deaths in 2008, ranking the fifth most frequently diagnosed cancer and the third leading cause of cancer death [19]. Admittedly, early diagnosis is of laudability and benefit for long-term survival of GC, but it is extremely challenging. Therefore, exploring new predictive biomarkers are urgently demanded for early detection, progression and prognosis of GC. So far, a great deal of evidence has demonstrated that GSDMs dysregulation can influence the development of cancers [11–13], but further bioinformatics analysis of GC has yet to be performed. In the current study, via public resource databases, we explored the mRNA expression and prognostic values of GSDMs in GC, which showed different results in the mRNA expression of GSDMs in GC. In TIMER database, GSDMB, GSDMC, GSDMD, GSDME were with higher expression in STAD than normal tissue, while via UALCAN database, GSDMA, GSDMB, GSDMD and GSDME were higher in STAD than in normal tissues. GSDMB, GSDMD and GSDME showed the highly consistent trend deducing potential role in tumorigenesis. Furthermore, tumor stage and tumor grade were analyzed in the expression of GSDMs. The results of GSDMs expression in different tumor stages suggested that advanced tumor stage existed distinct mechanisms blocking the expression of GSDMs. From the protein expression level, GSDMA and GSDMD showed consistent results, which were highly expressed in STAD. The accordant trend suggested that GSDMs perhaps played the role in oncogenesis and progression. To appraise the effect of GSDMs in oncogenesis of STAD, the prognostic assessment was performed. The GSDME gave the accordant result showing the prognostic value in STAD. Therefore, GSDME was paid our attention to evaluate the potential mechanism as a prediction biomarker. So far, many studies have reported that GSDME was involved in oncogenesis and chemoresistance. Knockdown GSDME could markedly suppress the growth of hepatocellular carcinoma [20]. GSDME-EGFR interaction involved the development of non-small cell lung cancer [21], which could open the horizon of cancer pathogenesis. By inducing caspase 3-GSDME pathway, CC-115 exerts antitumor effects in lung adenocarcinoma [22]. It is believed that GSDME could be a hopeful predictive and therapeutic marker of multiple tumors.

To further elucidate the underlying mechanism of GSDMs in GC tumorigenesis, progression and prognosis, we deliberated the biological function and immune infiltrates related to GSDMs expression. The molecular functions regarding GSDMs were focused on protease activity. It is reported that proteolytic networks could regulate tumor angiogenesis, invasion and signaling pathways in the tumor microenvironment involving chemokines, cytokines, and kinases [23]. Next, we analyzed the correlation between GSDMs expression and immune cell infiltration. Emerging evidence has indicated that immune microenvironment plays a key role in the GC tumorigenesis, progression and prognosis [24–27] and become a new determinant of immunotherapy response and clinical outcome [28–29]. It is said that tumor infiltrating lymphocytes (TILs) (including T cells, B cells and NK cells) were increased in STAD, especially among advanced cases, which suggest that TILs may be associated with tumor immune escape and with dysfunction of T cell in STAD [30]. Moreover, several types of gastric cancer showed immune tolerance, which infiltrated with high levels of TILs and low PD-L1 [31]. In our study, the expression level of GSDME was positive correlation with the CD4+ T cells, CD8+ T cells, neutrophils, dendritic cells and macrophages infiltration, suggesting that GSDME may be involved in the immunomodulatory mechanisms of STAD. Recently, it is recognized that the tumor microenvironment (TME) play an important role in enabling tumors to proliferate and metastasize, and single-cell RNA sequencing revealed that TME of GC was enriched for stromal cells, macrophages, dendritic cells and Tregs [32]. Oshi et al [33] reported GC with high angiogenesis score was significantly associated with a lower infiltration of Th1, Th2 cells, and dendritic cells, and a higher infiltration of M1 macrophages, and was also associated with shorter survival. To our best knowledge, tumor-associated macrophages (TAMs), a critical member of TME, participated in the tumorigenesis and development of GC [33–34]. Recent studies have indicated that TAMs can promote tumor progression via taking part in the immune regulation of GC [35–37]. Huo et al [38] confirmed the prognostic value of TAMs for GC and pointed out that GC patients with higher macrophage infiltration had a poor prognosis. It has been reported that elevated level of peripheral or intratumoral neutrophils were accompanied with poor GC patient survival [39–40], which suggested neutrophils play an important role in promoting pathological process of GC. Recent study illuminated pathogenic roles of neutrophils in GC with a novel mechanism that tumor tissue can attract neutrophils migration by CXCL6/CXCL8-CXCR1 interactions and lead to accumulation of neutrophils in GC tumors [41]. In these decades, there are different opinions about the role of tumor-infiltrating immune cells. It is well established that the immune cells are a double-blade sword, potentially promoting, as well as inhibiting, the development of cancer, which need further investigate [42–43]. The present study indicated that there were positive relationships between GSDME expression level and infiltration level of immune cells, suggesting GSDME might play a critical role in the regulation of immune infiltrating cells in GC.

## Conclusion

Owing to the critical role of GSDMs in pyroptosis and tumor progression, we systematically evaluated the expression levels and prognostic value of GSDMs in GC. Our study demonstrated that GSDME might be a reliable diagnostic and prognostic biomarker and associated with oncogenic signaling pathways and immune in GC. However, our study can only provide a preliminary theoretical basis and needs to be further verified by follow-up studies.

## Data Availability

All relevant data are within the manuscript and its Supporting Information files

## Competing interests

The authors declare that they have no competing interests.

## Acknowledgements

This project was supported by a grant from Changshu science and technology project (CS202018,CSWZD202043, CSWS202106).

## Notes

### Competing Interest Statement

The authors have declared no competing interest.

### Funding Statement

The author(s) received no specific funding for this work.

### Author Declarations

Ethics committee of Changshu Medicine Examination Insitute

